## Supplementary material for "Evaluating Modes of Influenza Transmission (EMIT-2): Insights from a Controlled Human Influenza Virus Infection Transmission Trial (CHIVITT)": S1 Fig. Consort diagram for Recipients.docx

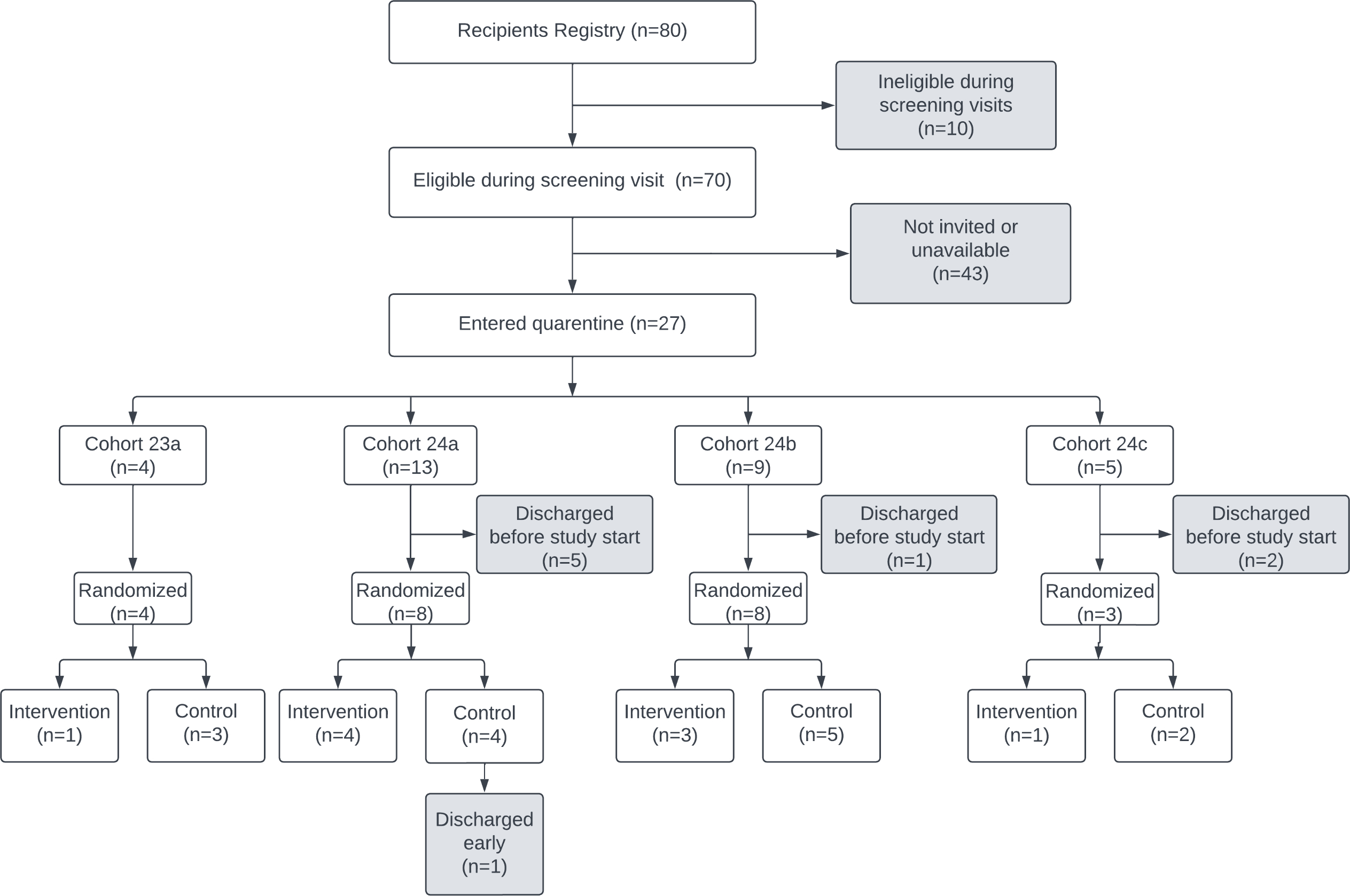

### S1 Fig. Consort diagram for Recipients

Three Recipients from Cohort 23a re-enrolled in 2024. Two initially joined Cohort 24a, but one was discharged before the study began due to a respiratory infection other than influenza and later re-enrolled in Cohort 24c. Another Recipient from Cohort 23a also joined Cohort 24c. As a result, the total number of distinct individuals who entered quarantine was 27, rather than the summed total (31) from each individual cohort.

Note: Influenza Donors were only enrolled in cohorts 24b and 24c; no influenza Donors were enrolled in 23a or 24a.
