## Supplementary material for "Evaluating Modes of Influenza Transmission (EMIT-2): Insights from a Controlled Human Influenza Virus Infection Transmission Trial (CHIVITT)": S1 Table. All Recipients demographics .docx

|  | Cohort 23a | | Cohort 24a | | Cohort 24b | | Cohort 24c | | All |
| --- | --- | --- | --- | --- | --- | --- | --- | --- | --- |
|  | **CTL** | **INT** | **CTL** | **INT** | **CTL** | **INT** | **CTL** | **INT** |  |
| Number of participants | 3 | 1 | 4 | 4 | 5 | 3 | 2 | 1 | 23 |
| Female, N (%) | 1 (33) | 0 (0) | 1 (25) | 1 (25) | 2 (40) | 2 (67) | 1 (50) | 1 (100) | 9 (39) |
| Age, mean (SD) | 38.3 (1.53) | 44.0 (-) | 39.3 (5.32) | 42.0 (8.04) | 36.4 (10.5) | 29.0 (6.08) | 41.0 (4.24) | 39.0 (-) | 38.0 (7.37) |
| Vaccinated^a^, N (%) | 1 (33) | 0 (0) | 2 (50) | 2 (50) | 1 (20) | 1 (33) | 0 (0) | 0 (0) | 7 (30) |
| Latino, N (%) | 0 (0) | 0 (0) | 0 (0) | 1 (25) | 0 (0) | 1 (33) | 0 (0) | 0 (0) | 2 (9) |
| Race |  |  |  |  |  |  |  |  |  |
| Asian, N (%) | 0 (0) | 1 (100) | 1 (25) | 0 (0) | 0 (0) | 0 (0) | 0 (0) | 0 (0) | 2 (9) |
| Black or African American, N (%) | 0 (0) | 0 (0) | 1 (25) | 2 (50) | 2 (40) | 1 (33) | 1 (50) | 0 (0) | 7 (30) |
| White, N (%) | 3 (100) | 0 (0) | 2 (50) | 1 (25) | 3 (60) | 2 (67) | 1 (50) | 1 (100) | 13 (56) |
| Other races, N (%) | 0 (0) | 0 (0) | 0 (0) | 1 (25) | 0 (0) | 0 (0) | 0 (0) | 0 (0) | 1 (4) |

a. Vaccinated against influenza in the past 6 months at the time of entering the quarantine facility.
