## Supplementary material for "Evaluating Modes of Influenza Transmission (EMIT-2): Insights from a Controlled Human Influenza Virus Infection Transmission Trial (CHIVITT)": S1 Text. EMIT-2 Study Team.docx

### **S1 Text. EMIT-2 Study Team (Alphabetical by Last Name)**

Shuo Chen

Wilbur H. Chen

Kristen K. Coleman

Benjamin J. Cowling

Don L. DeVoe

Gregg Duncan

Arantza Eiguren-Fernandez

Yi Esparza

Jennifer German

Aubree Gordon

Filbert Hong

Yoshihiro Kawaoka

Florian Krammer

Jianyu Lai

Gregory S. Lewis

Kathleen M. McPhaul

Donald K. Milton

Gabriele Neumann

Justin R. Ortiz

Margaret Scull

Jelena Srebric

Isabel Sierra Maldonado

S.-H. Sheldon Tai

Shengwei Zhu
