## Supplementary material for "Evaluating Modes of Influenza Transmission (EMIT-2): Insights from a Controlled Human Influenza Virus Infection Transmission Trial (CHIVITT)": S2 Fig. Consort diagram for Donors.docx

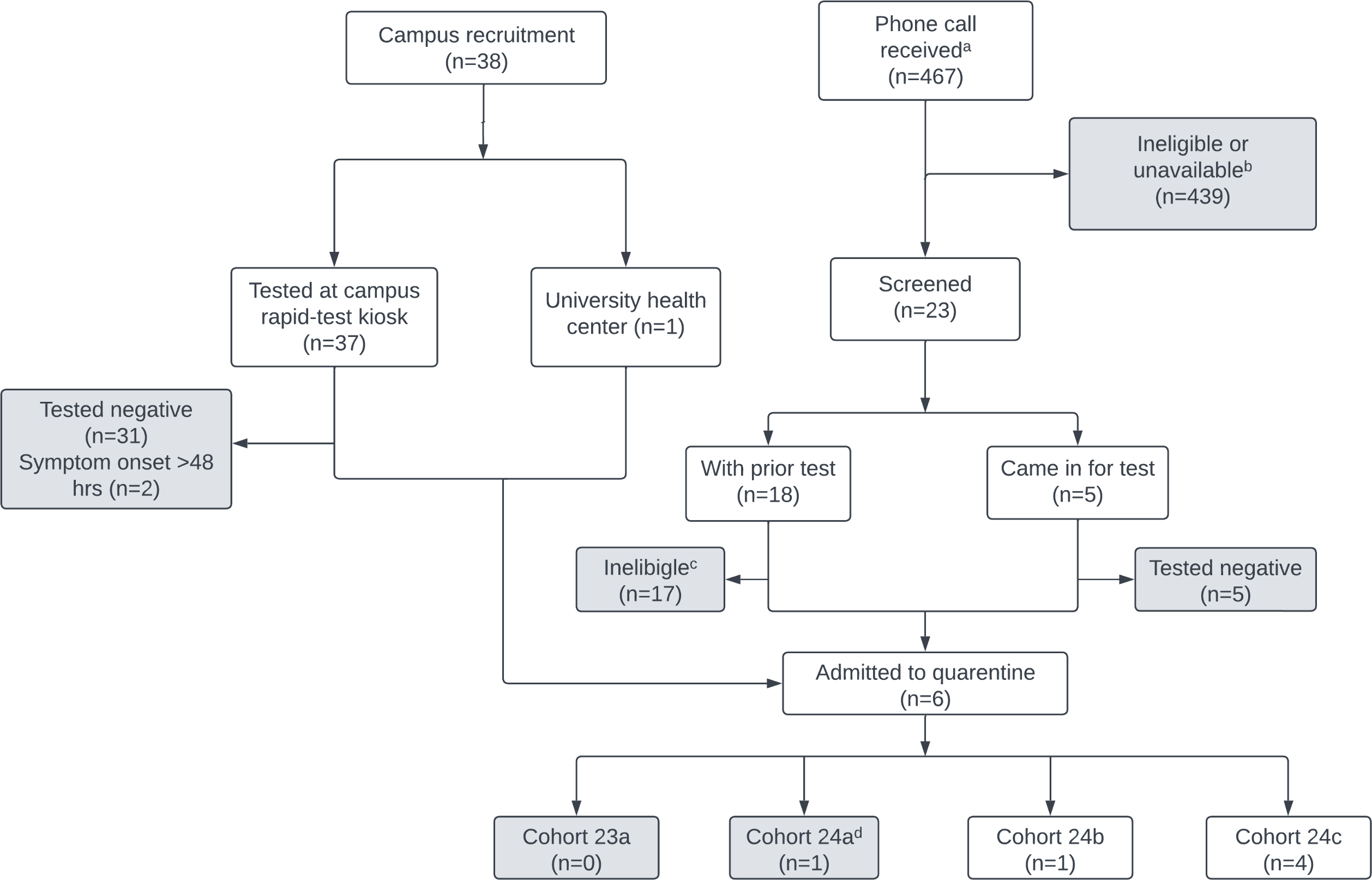

### S2 Fig. Consort diagram for Donors

a. “Phone call received” includes inquiries from social media, flyers, television, and other advertisements.

b. Many individuals were ineligible due to lack of influenza infection or were unavailable to commit to the quarantine period.

c. Ineligibility among screened individuals included symptom onset >48 hours or underlying medical conditions that did not meet eligibility criteria (S3 Text).

d. The Donor enrolled in cohort 24a, recruited from among the phone calls received, tested positive for a seasonal coronavirus (229E); no Donors were recruited for cohort 23a.
