## Supplementary material for "Evaluating Modes of Influenza Transmission (EMIT-2): Insights from a Controlled Human Influenza Virus Infection Transmission Trial (CHIVITT)": S2 Table. All Donors demographics.docx

|  | **Cohort 24a** | **Cohort 24b** | **Cohort 24c** | **All** |
| --- | --- | --- | --- | --- |
| **Number of participants** | 1 | 1 | 4 | 6 |
| **Female, N (%)** | 1 (100) | 1 (100) | 3 (75) | 5 (83) |
| **Age, mean (SD)** | 25.0 (-) | 23.0 (-) | 20.5 (0.577) | 21.7 (1.97) |
| **Vaccination, N (%)** | 0 (0) | 0 (0) | 2 (50) | 2 (33) |
| **Latino, N (%)** | 0 (0) | 0 (0) | 2 (50) | 2 (33) |
| **Race** |  |  |  |  |
| **Asian, N (%)** | 0 (0) | 1 (100) | 0 (0) | 1 (17) |
| **Black or African American, N (%)** | 1 (100) | 0 (0) | 2 (50) | 3 (50) |
| **White, N (%)** | 0 (0) | 0 (0) | 2 (50) | 2 (33) |
