## Supplementary material for "Evaluating Modes of Influenza Transmission (EMIT-2): Insights from a Controlled Human Influenza Virus Infection Transmission Trial (CHIVITT)": S2 Text. Acknowledged Contributors.docx

### **S2 Text. Acknowledged Contributors (Alphabetical by Last Name)**

**Public Health AeroBiology Lab at the University of Maryland, College Park**

Tiffany Akotia, Beata Assadi, Julianna Corrigan, Esther Fadaka, Naja Fadul, T. Louie Gold, Grace Herron, Zahra Iskandar, Petri Kalliomäki, Mariana Lim, Molly Oertel, Veerapetch Petchgar, Anna Pulley, Sulakkhana De Saram, Maria Schanz, Alycia Smith, William Smith, Aditya Srikakulapu, Rachel Tackett, Candela Vazquez, Olga Volchansky, and Jonathan Vyskocil.

**Maryland MEMS & Microfluidics Lab at the University of Maryland, College Park**

Proma Bhattacharaya, Siddarth Raghu Srimathi

**Duncan Lab at the University of Maryland, College Park**

Shadin Doski

**The Center for Sustainability in the Built Environment (City@UMD) at the University of Maryland, College Park**

Pranav Shinde, Gautam Vanama

**University of Maryland, Baltimore**

Karthikeyan Appadurai, Saleha Bharde, Colleen Boyce, Meagan Deming, Samantha Fletcher, Shirley George, Andrew Gilmore, Nancy Greenberg, Sophie Harper, Sarah Hart, Susan Holian, Rebecca Hudson, Alexander Karasin, Alyson Kwon, Moishe Lerner, Sarah Litts, Tadashi Maemura, Melissa Marini, Kaitlin Mason, Suemoal Mathew, Ifayet Mayo, Sherry McCammon, Megan McGilvray, Julieta Nacario, Rosary Necesario, Kathleen Neuzil, Candice Onyeama, Jennifer Oshinsky, Lynnee Roane, Ligia Sandor, Barbara Sinagra, Jody Suliman, Constance Thomas, Jennifer Winkler.
