## Supplementary material for "Evaluating Modes of Influenza Transmission (EMIT-2): Insights from a Controlled Human Influenza Virus Infection Transmission Trial (CHIVITT)": S3 Fig. Lack of correlation of Donor exhaled breath aerosol viral RNA with viral RNA in either Donor MTS or saliva samples.docx

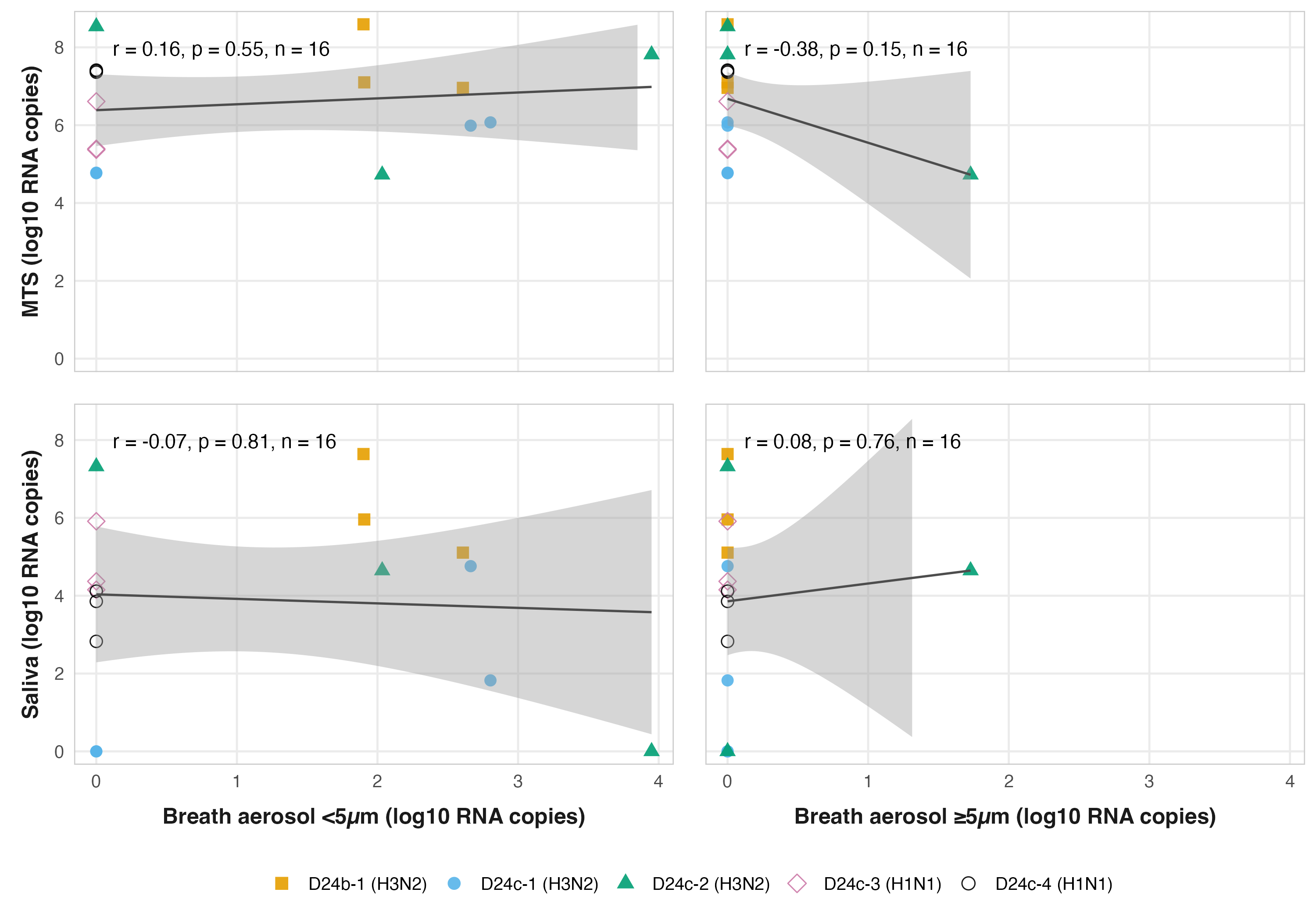

### S3 Fig. Lack of correlation of Donor exhaled breath aerosol viral RNA with viral RNA in either Donor MTS or saliva samples.

Scatterplots show pairwise correlations of viral RNA concentrations measured from exhaled breath aerosol samples (left column: particles <5 µm; right column: particles ≥5 µm) against matched MTS (top row) and saliva (bottom row) samples collected from the same Donors on the same days. Each point represents one Donor–day measurement, with colors and shapes denoting individual Donors. Solid points represent samples from Donors with H3N2 infections, while hollow points represent samples from Donors with H1N1 infections. Black lines indicate linear regression fits with 95% confidence intervals. Pearson correlation coefficients (r) and sample sizes (n) are shown within each plot.
