## Supplementary material for "Evaluating Modes of Influenza Transmission (EMIT-2): Insights from a Controlled Human Influenza Virus Infection Transmission Trial (CHIVITT)": S3 Table. Exposure events & activities.docx

| Cohort | Date (Cohort Day) | Event | Activities | Object passed & swabbed | Recipient Number | Donor Number | Staff Number | Duration (Minutes) |
| --- | --- | --- | --- | --- | --- | --- | --- | --- |
| Cohort 24b | 2024-01-30 (7) | 1 | Icebreaker questions;  Telestrations;  Movie/TV | Marker | 8 | 1 | 2 | 250 |
|  | 2024-01-31 (8) | 2 | Yoga/stretch;  Discussion/chat;  Heads Up!;  UNO | Tablet computer | 8 | 1 | 2 | 120 |
|  |  | 3 | Discussion/chat;  Puzzles;  Karaoke | Marker | 8 | 1 | 2 | 240 |
|  |  | 4 | Movie/TV;  Telestrations;  Discussion/chat | Marker | 8 | 1 | 2 | 236 |
|  | 2024-02-01 (9) | 5 | Stretching;  Just Dance;  Blank Slate | Marker | 8 | 1 | 2 | 120 |
|  |  | 6 | Pantone;  Playing cards (Poker) | Marker | 8 | 1 | 2 | 205 |
| Cohort 24c  Window 1 | 2024-02-16 (3) | 1 | Icebreaker questions;  Movie/TV | Marker | 3 | 1 | 2 | 111 |
|  |  | 2 | Movie/TV;  Blank Slate;  Discussion/chat | Marker | 3 | 1 | 2 | 207 |
|  | 2024-02-17 (4) | 3 | Stretching;  Movie/TV | Marker | 3 | 1 | 2 | 128 |
|  |  | 4 | Blank Slate;  Secret Hitler;  Codenames | Marker | 3 | 1 | 2 | 239 |
|  |  | 5 | UNO;  Pantone;  Cards Against Humanity;  Movie/TV;  Icebreaker questions | Marker | 3 | 2 | 2 | 240 |
|  | 2024-02-18  (5) | 6 | Stretching;  Discussion/chat;  Puzzles;  Coloring | Marker | 3 | 2 | 2 | 237 |
|  |  | 7 | Coloring;  Secret Hitler;  UNO;  Blank Slate | Marker | 3 | 2 | 2 | 233 |
|  |  | 8 | Telestrations;  Cards Against Humanity;  Movie/TV | Marker | 3 | 2 | 2 | 233 |
|  | 2024-02-19  (6) | 9 | Stretching; Discussion/chat;  GeoGuessr;  Spyfall;  Online escape room;  Coup | Marker | 3 | 2 | 2 | 236 |
|  |  | 10 | Spoons;  BS(card game);  Wordle;  Trivia | Marker | 3 | 2 | 2 | 231 |
|  |  | 11 | UNO;  Coloring;  Discussion/chat;  Movie/TV | Marker | 3 | 1 | 2 | 228 |
| Cohort 24c  Window 2 | 2024-02-21  (8) | 1 | Icebreaker questions;  Blank Slate;  Discussion/chat;  Cards Against Humanity;  Movie/TV | Marker | 3 | 2 | 2 | 242 |
|  | 2024-02-22  (9) | 2 | Yoga/stretch;  Codenames;  UNO;  Telestrations | Marker | 3 | 2 | 2 | 240 |
|  |  | 3 | Heads Up!;  Hangman;  UNO | Tablet computer | 3 | 2 | 2 | 243 |
|  |  | 4 | Secret Hitler;  Puzzles;  Coup;  Movie/TV | Microphone | 3 | 2 | 2 | 240 |
|  | 2024-02-23  (10) | 5 | Discussion/chat;  Online Jeopardy;  Guess Who;  UNO | Marker | 3 | 2 | 2 | 234 |
|  |  | 6 | Murder Mystery;  Spoons;  Playing cards (Poker);  Coloring;  Movie/TV | Microphone | 3 | 2 | 2 | 240 |
