## Supplementary material for "Evaluating Modes of Influenza Transmission (EMIT-2): Insights from a Controlled Human Influenza Virus Infection Transmission Trial (CHIVITT)": S3 Text. Eligibility Criteria.docx

### **Part I: Donor Eligibility Criteria**

#### Donor Inclusion Criteria

1. Provides written informed consent, able to comply with the planned study procedures, available for between 2 and 5 days stay in the research quarantine unit for the CHIVITT, and have the ability to attend the scheduled follow-up visits.
2. Subjects must be able to comprehend the study requirements, as evidenced by a score of ≥70% or better on the comprehension assessment (two attempts permitted).
3. Males and non-pregnant, non-breastfeeding females^1^ aged ≥18 and ≤59 years of age, at time of initial consent.

^1^Pregnancy and breastfeeding status to be determined by self-report

4. Laboratory-confirmed influenza infection^2^ within the past 48 hours at time of entry into the exposure event.

^2^A rapid antigen test in the setting of known local influenza activity and with symptoms suggestive of influenza at that time is acceptable

5. Within the past 48 hours at time of entry into the exposure event, onset of influenza-like illness, *defined as fever* (measured oral temperature of ≥100.2°F or self-reported fever in the absence of a measured temperature) AND cough or sore throat, or onset of less specific symptoms with a positive molecular test for influenza virus infection.*.*
6. No self-reported or known history of alcohol or drug abuse within the past two years and no illicit drug use within the last 30 days.
7. Do not have clinically significant medical, psychiatric, and chronic or intermittent health conditions including those listed in Exclusion Criteria*.*
8. Does not have an ongoing symptomatic condition^3^ for which subject has had or has ongoing medical investigations but has not yet received a diagnosis or treatment plan.

^3^e.g., ongoing and debilitating fatigue without a diagnosis for the symptom.

9. Agrees to the collection of specimens for secondary research.

#### Donor Exclusion Criteria

1. Female of childbearing potential who is breastfeeding or has positive urine pregnancy test upon admission to the hotel quarantine unit.
2. Presence of self-reported or medically documented significant medical or psychiatric condition(s)^5^

^5^Significant medical or psychiatric conditions include but are not limited to:

a. Respiratory disease (e.g., chronic obstructive pulmonary disease [COPD], asthma, cystic fibrosis) requiring daily medications^6^ currently or any treatment of respiratory disease exacerbations or hospitalizations for acute respiratory illnesses (e.g., asthma exacerbation) in the last 5 years.

^6^ Asthma medications: inhaled, oral, or intravenous (IV) corticosteroids, leukotriene modifiers, long and short acting beta agonists, theophylline, ipratropium, biologics.

b. Significant cardiovascular disease (e.g., congestive heart failure, cardiomyopathy, ischemic heart disease) or history of myocarditis or pericarditis as an adult.
c. Neurological or neurodevelopmental conditions (e.g., epilepsy, stroke, seizures, encephalopathy, focal neurologic deficits, Guillain-Barré syndrome, encephalomyelitis or transverse myelitis).
d. Ongoing malignancy or recent diagnosis of malignancy, including leukemia; treated, non melanoma skin cancers are permissible.
e. An autoimmune disease.
f. An immunodeficiency of any cause.
g. A blood disorder (e.g., sickle cell disease)
h. Endocrine disorders (e.g., diabetes)
i. Liver, kidney, metabolic disorders
j. BMI ≥40 kg/m^2^
k. Any other condition or behavior that in the opinion of the PI would affect the ability to participate in the transmission study over the next several days.

3. Presence of immunosuppression or any medications that may be associated with impaired immune responsiveness^7^.

^7^Including, but not limited to, corticosteroids exceeding 10 mg/day of prednisone equivalent, immunoglobulin, interferon, immunomodulators, cytotoxic drugs, or systemic corticosteroids or other similar or toxic drugs during the preceding 12-month period. Low dose topical and intranasal steroid preparations used for a discrete period are permitted.

4. Is a habitual smoker^8^ of tobacco, marijuana, or e-cigarettes per self-report.

^8^Habitual smokers are those who smoke or vape more than four cigarettes, other tobacco products, e-cigarettes or marijuana in a week for more than three months or use an inhaled nicotine or marijuana product more than 3 days a week on average. Edible or patch forms of tobacco or marijuana products do not constitute an exclusion.

5. Known allergy or intolerance to treatments for influenza and other respiratory infections (including but not limited to acetaminophen/paracetamol).
6. History of a previous severe allergic reaction to medicines of any kind with generalized urticaria, angioedema, or anaphylaxis.
7. Presence of co-infection with SARS-CoV-2, as detected via a multiplex nucleic acid amplification test (e.g., Biofire).
8. Participating in any other interventional clinical research study that has a scheduled intervention 30 days prior to the CHIVITT or 30 after discharge from the research quarantine unit.
9. Any condition, to include medical and psychiatric conditions, that in the opinion of the Investigator, might interfere with the safety of the subject or the study objectives.

### **Part II: Recipient Eligibility Criteria**

#### Recipient Inclusion Criteria

1. Enrolled in the Recipient Protocol (UMB IRB HP-97730)
2. Provides written informed consent, able to comply with the planned study procedures, be available for an up to ~14-day stay for the CHIVITT and have the ability to attend the scheduled follow-up visits.
3. Subjects must be able to comprehend the study requirements, as evidenced by a score of ≥70% or better on the comprehension assessment (two attempts permitted).
4. No significant change (for the worse) in general health history or in concomitant medication use, as compared from their responses collected during screening (EMIT-2 Recipient Protocol).
5. Agree not to meet with other participants (recipients or donors) outside of the programmed exposure events during the course of their participation in the CHIVITT.

#### Recipient Exclusion Criteria

1. Female of childbearing potential who has a positive urine pregnancy test within 24 hours of admission to the hotel quarantine unit or is breastfeeding or planning to become pregnant within 2 months after entry into a CHIVITT.
2. Presence of infection with influenza, SARS-CoV-2, or other respiratory pathogens detected via a multiplex nucleic acid amplification test (e.g., Biofire) at admission to the hotel quarantine facility.
3. Within the past 72 hours, presence of influenza-like illness, as defined as fever of ≥100.2°F AND cough or sore throat, in the absence of an alternative cause.
4. Receipt of any blood products within the past 2 months.
5. Does not agree to provide permission for secondary research use of extra samples collected and stored specimens.
6. Habitual smoker of tobacco, marijuana, or e-cigarettes per self-report. (Habitual smokers are those who smoke or vape more than four cigarettes, other tobacco products, e-cigarettes or marijuana in a week for more than three months or use an inhaled nicotine or marijuana product more than 3 days a week on average. Edible or patch forms of tobacco or marijuana products do not constitute an exclusion.)
7. Self-reported or known history of alcohol or drug abuse in the past two years and/or illicit drug use within the last 30 days. (*Prescribed stimulants for the treatment of ADHD and cannabinoids use do not constitute exclusionary criteria*)
8. Has an ongoing symptomatic condition^1^ for which the subject has had or has ongoing medical investigations but has not yet received a diagnosis or treatment plan.

^1^e.g., ongoing chronic fatigue without a diagnosis for symptom.

9. Presence of self-reported or medically documented significant medical or psychiatric condition(s)^2^

^2^Significant medical or psychiatric conditions include but are not limited to:

a. Respiratory disease (e.g., chronic obstructive pulmonary disease [COPD], asthma, cystic fibrosis) requiring daily medications* currently or any treatment of respiratory disease exacerbations or hospitalizations for acute respiratory illnesses (e.g., asthma exacerbation) in the last 5 years.

* Asthma medications: inhaled, oral, or intravenous (IV) corticosteroids, leukotriene modifiers, long and short acting beta agonists, theophylline, ipratropium, biologics.

b. Significant cardiovascular disease (e.g., congestive heart failure, cardiomyopathy, ischemic heart disease) or history of myocarditis or pericarditis as an adult.
c. Neurological or neurodevelopmental conditions (e.g., epilepsy, stroke, seizures, encephalopathy, focal neurologic deficits, Guillain-Barré syndrome, encephalomyelitis or transverse myelitis).
d. Ongoing malignancy or recent diagnosis of malignancy, including leukemia; treated, non-melanoma skin cancers are permissible.
e. An autoimmune disease.
f. An immunodeficiency of any cause.
g. A blood disorder (e.g., sickle cell disease)
h. Endocrine disorders (e.g., diabetes)
i. Liver, kidney, metabolic disorders
j. BMI ≥40 kg/m2
k. Any other condition or behavior that in the opinion of the PI would affect the ability to participate in the screening or future transmission studies.
10. Presence of immunosuppression or any medications that may be associated with impaired immune responsiveness^3^.

^3^Including, but not limited to, corticosteroids exceeding 10 mg/day of prednisone equivalent, immunoglobulin, interferon, immunomodulators, cytotoxic drugs, or systemic corticosteroids or other similar or toxic drugs during the preceding 12-month period. Low dose topical and intranasal steroid preparations used for a discrete period are permitted.

11. Known allergy or intolerance to treatments for influenza and other respiratory infections (including but not limited to oseltamivir, baloxavir, acetaminophen/paracetamol).
12. History of a previous severe allergic reaction to medicines of any kind with generalized urticaria, angioedema, or anaphylaxis.
13. Participating in any other interventional clinical trial that has a scheduled intervention 30 days prior to the start of the CHIVITT or 30 after discharge from the research quarantine unit.
