## Supplementary material for "Evaluating Modes of Influenza Transmission (EMIT-2): Insights from a Controlled Human Influenza Virus Infection Transmission Trial (CHIVITT)": S4 Fig. Viral load in ambient bioaerosol and surface swab samples.docx

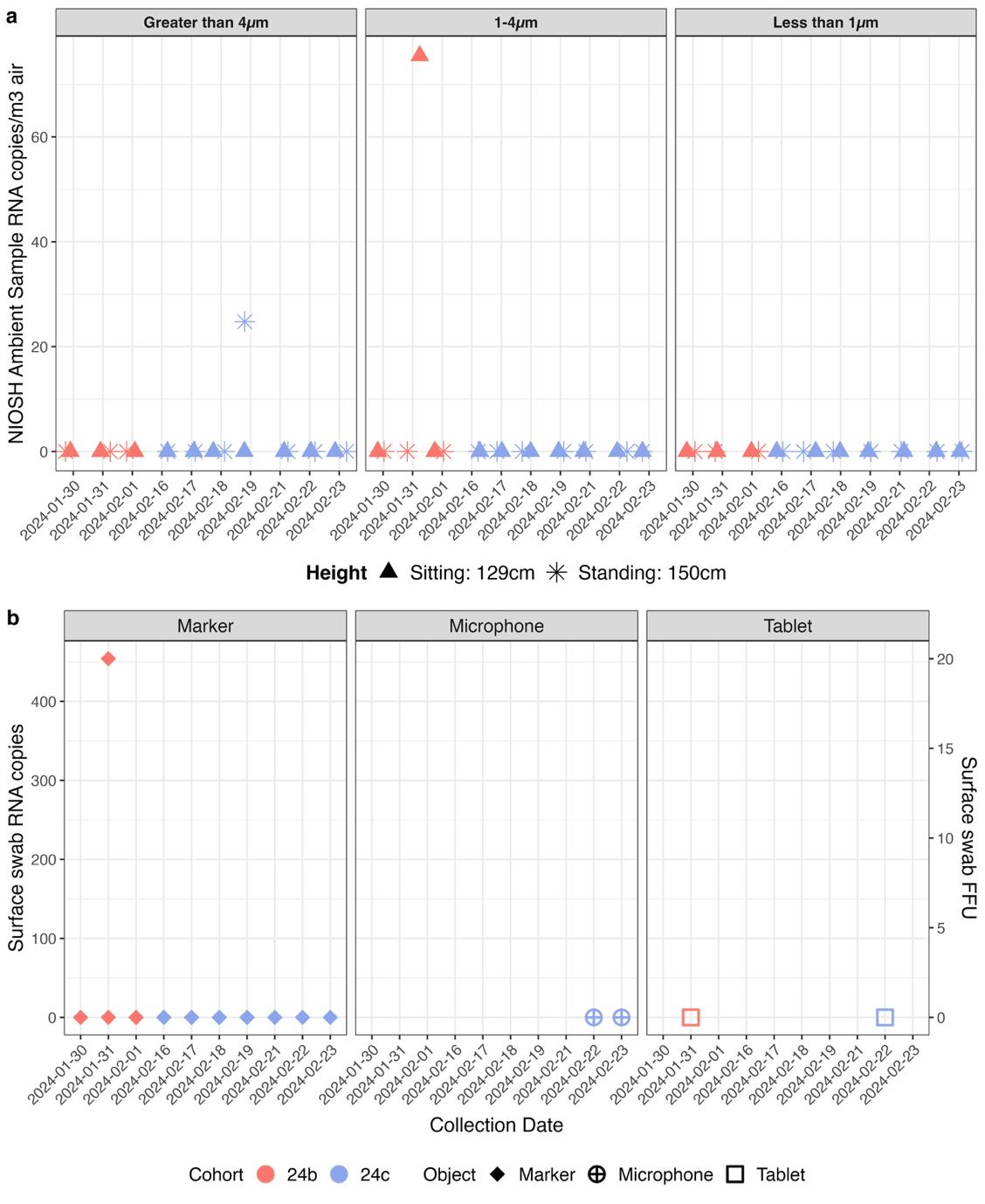

### S4 Fig. Viral load in ambient bioaerosol and surface swab samples

(a) RNA copies from ambient bioaerosol samples collected using NIOSH BC-251 bioaerosol sampling devices, stratified by size fraction (>4 μm, 1–4 μm, and <1 μm).

(b) RNA copies (left y-axis) and infectious viral load (FFU, right y-axis) from surface swabs collected from marker pens, microphones, and tablets. Each point represents one sample. Only one surface sample, collected from a marker pen, contained virus detected by dPCR; it was also positive by viral culture.
