## Supplementary material for "Evaluating Modes of Influenza Transmission (EMIT-2): Insights from a Controlled Human Influenza Virus Infection Transmission Trial (CHIVITT)": S4 Table. Summary of Donors cough and sneeze counts during 30-minute breath sample sampling.docx

| Donor | Cough counts,  median(min, max) | Sneeze counts,  median(min, max) |
| --- | --- | --- |
| D24b-1 | 0 (0, 6) | 0 (0, 0) |
| D24c-1 | 0 (0, 0) | 0 (0, 0) |
| D24c-2 | 0 (0, 6) | 0 (0, 0) |
| D24c-3 | 0 (0, 4) | 0 (0, 1) |
| D24c-4 | 5 (0, 16) | 0 (0, 0) |
| All | 0 (0, 16) | 0 (0, 1) |
