## Supplementary material for "Evaluating Modes of Influenza Transmission (EMIT-2): Insights from a Controlled Human Influenza Virus Infection Transmission Trial (CHIVITT)": S4 Text. Supplementary Methods.docx

### Power Analysis

Our original power analysis was based on the assumptions that: 1) SAR without interventions is 25%; 2) viral aerosol inhalation attributes to 75% of influenza transmission; 3) half of the aerosol inhalation is due to short-range aerosol exposure, and another half is attributable to long-range aerosol exposure. Having 18 Recipients per group and 3 pairs of quarantine cohorts where one cohort’s exposures occurred with and the other without air sanitation (Total = 108 Recipients, with 54 per arm), we would have 78% power to detect 74% reduction in SAR (from 21.9% to 5.9%).

### Randomization

For each sampled exposure event (EE), we randomly selected one control Recipient (CR) and one Intervention Recipient (IR) to wear a sampling kit. To select these participants, we used a script in R that performs a lottery once a day, via crontab module. The script then posted the results to a de-identified webpage for use by the study team.

### Sample Preparation

G-II exhaled breath aerosol samples were processed as previously described [1]. Mid-turbinate swabs and surface swabs were eluted in 3 mL and 1 mL Viral Transport Media (BD), respectively.

Ambient air samples were processed by adding 0.4 mL Dulbecco’s phosphate buffered saline (DPBS) containing 0.1% bovine serum albumin (BSA) and 0.6 mL Lysis/Binding Solution Concentrate (Applied Biosystems) to each stage of the NIOSH samples prior to vortex mixing and aliquoting.

Aliquots of the samples prepared for fluorescence focus assay were stored at 4°C until the assay was set up within 24 hours. The other aliquots were stored at -80°C until further analyses.

### Digital PCR (dPCR)

We quantified influenza A virus RNA using the QIAcuity One dPCR System (Qiagen). Nucleic acids from G-II, mid-turbinate swab, saliva, surface swab, and ambient air samples were first extracted using the MagMAX Pathogen RNA/DNA Kit (Applied Biosystems) and the KingFisher Duo Prime Purification System (Thermo Scientific), following the manufacturers’ instructions. For each sample, we prepared 40 µL dPCR reaction mix consisted of 20 µL of freshly extracted nucleic acids, 1 × OneStep Advanced Probe Master Mix and OneStep Advanced RT Mix from the QIAcuity OneStep Advanced Probe Kit (Qiagen), recently updated influenza A virus matrix (M) gene segment primers Forward 1 (400 nM), Forward 2 (400 nM), Reverse 1 (600 nM), Reverse 2 (200 nM), and 200 nM Probe (described in [2] and synthesized by Integrated DNA Technologies). After thorough mixing, the dPCR mixes were transferred into QIAcuity Nanoplates with 26k partitions per well (Qiagen) and sealed. After distributing the reaction mix into Nanoplate partitions, the QIAcuity One dPCR System incubated each plate at 50°C 40 min for reverse transcription and cycled through 95°C 5 sec denaturation and 55°C 30 sec annealing/extension 45 times. At the end of thermal cycling, the dPCR system calculates copy numbers in the reactions and 95% confidence intervals (CI) based on numbers of positive and negative partitions, which were back calculated to copy numbers in original samples. We tested samples with high viral loads in multiple dilutions to ensure that at least one of them was within the dynamic range of the dPCR assay. Samples that had ≤ 1 positive partition were considered negative in subsequent analyses.

### Focus Assay

To measure the infectivity of seasonal influenza A viruses in G-II, mid-turbinate swab, and surface swab samples collected in this study, we performed fluorescence focus assays using the humanized Madin-Darby canine kidney (hCK) cells kindly provided by Dr. Yoshihiro Kawaoka [3] and adapted a previously published method [1]. Briefly, hCK monolayers in 96-well plates were washed with DPBS to remove fetal bovine serum (FBS) in the media and incubated with samples for 1 h at 37°C in the presence of 1.5-2 µg/mL TPCK-treated trypsin (Sigma-Aldrich). Following virus adsorption and entry, media containing FBS were added to the culture to inhibit multiple rounds of viral replication by inactivating extracellular trypsin. Inoculated monolayers were incubated at 37°C for a total of 8 h before they were fixed with formalin (Fisher Scientific). We subsequently identified infected cells by immunofluorescence staining. First the cells were permeabilized with 0.25% Tween 20 (Sigma-Aldrich) and non-specific antibody bindings blocked with 1% BSA. Influenza A virus nucleoprotein was stained using AA5h primary antibody (Santa Cruz Biotechnology) and AlexaFluor 488-conjugated goat-anti-mouse secondary antibody (Jackson ImmunoResearch Laboratories). Cell nucleus was visualized with 4',6-diamidino-2-phenylindole (Santa Cruz Biotechnology) staining. Positive cells were counted as focus-forming units (FFUs) using fluorescence microscopy, which were used to deduce FFUs in the original samples.

### Hemagglutination Inhibition Assay (HAI)

To remove non-specific virus inhibitors, 25 µL sera samples were treated with 75 µL receptor-destroying enzyme II (Denka Seiken) for 16 hours at 37ºC. Treatment was stopped with the addition of 75 µL 2.5% sodium citrate solution. After 1 hour incubation at 56ºC, 75 µL PBS was added to the samples, bringing the starting dilution to 1:10. Using 96-well V-bottom plates (Thermo-Scientific), samples were diluted two-fold in 25 µL/well PBS until a final dilution of 1:10,240. Virus was diluted to 8 hemagglutination activity units/50 μL in PBS, as determined previously by a hemagglutination assay. 25 μL/well of virus was added to the diluted sera plates, then plates were incubated for 30 minutes on a shaker at room temperature. After 30 minutes, 0.5% red blood cells from either turkey (for A/Victoria/22) or guinea pig (for A/Darwin/21 and B/Austria/21) were added to the plates at a volume of 50 μL/well. Plates were incubated at 4ºC for 1 hour (turkey blood) or 2 hours (guinea pig blood), until pellets were observed on the bottom of the blank wells. Titers were calculated by determining the last dilution at which a clear pellet was observed.

### Enzyme-Linked Immunosorbent Assay (ELISA)

An enzyme-linked immunosorbent assay (ELISA) was used to assess the area under the curve (AUC) for binding antibody responses against A/Victoria/4897/22 H1 and A/Darwin/6/21 H3. 96 well plates (Immulon 4 HBX; Thermo Fisher Scientific) were coated with recombinant HA protein diluted in phosphate-buffered saline (PBS) to a concentration of 2 μg/ml (50 μl/well) and incubated overnight at 4ºC. The next day, plates were washed with 1X PBS containing 0.1% Tween 20 (PBS-T). Plates were blocked with 200 μl/well of 3% nonfat dry milk powder (bioWORLD) in PBS-T at room temperature. After 1 hour, blocking solution was removed and plates were filled with 1% milk powder in PBS-T (dilution buffer). Sera samples were added at an initial dilution of 1:100 and serially diluted 3 fold to a final dilution of 1: 218,700. After incubating for 2 hours at room temperature, plates were washed three times with PBS-T. Fab-specific anti-human secondary antibody conjugated to horseradish peroxidase (HRP) was diluted 1:9000 in dilution buffer and added to the plates at 50 μl/well. Plates were incubated for 1 hour at room temperature and then washed four times with PBS-T. 100 µL/well of O-phenylenediamine dihydrochloride substrate (Sigma-Aldrich) was added to the plates. The reaction was stopped after 10 minutes by the addition of 50 µL/well of 3M HCl (Thermo Fisher Scientific). Optical density at 490 nm was measured using a Synergy 4 (BioTek) microplate reader. AUC values were calculated using GraphPad Prism 9.

### Computation of Maximal Inhalation Dose

We computed maximal exposure from ambient sampler measurements of RNA copies per cubic meter and, based on Recipient activities during exposure events an estimated average minute-ventilation rate of 16 L/min (~1 m^3^/hour). Using this estimate and the two positive NIOSH bioaerosol samples, we estimate that on January 31, Recipients in Cohort 24b were each exposed to at approximately 750 RNA copies (75 RNA copies/m^3^, for 10 hours of exposure events) via inhalation. On February 19, Recipients in Cohort 24c were exposed to 290 RNA copies (25 RNA copies/m^3^, for 11.6 hours of exposure events).

**References**

1.	Yan J, Grantham M, Pantelic J, et al. Infectious virus in exhaled breath of symptomatic seasonal influenza cases from a college community. Proc Natl Acad Sci USA **2018**; 115:1081–1086.

2.	Shu B, Kirby MK, Davis WG, et al. Multiplex Real-Time Reverse Transcription PCR for Influenza A Virus, Influenza B Virus, and Severe Acute Respiratory Syndrome Coronavirus 2. Emerg Infect Dis **2021**; 27:1821–1830.

3.	Takada K, Kawakami C, Fan S, et al. A humanized MDCK cell line for the efficient isolation and propagation of human influenza viruses. Nat Microbiol **2019**; 4:1268–1273.
