## Supplementary material for "Evaluating Modes of Influenza Transmission (EMIT-2): Insights from a Controlled Human Influenza Virus Infection Transmission Trial (CHIVITT)": S5 Fig. ELISA AUC 24b.docx

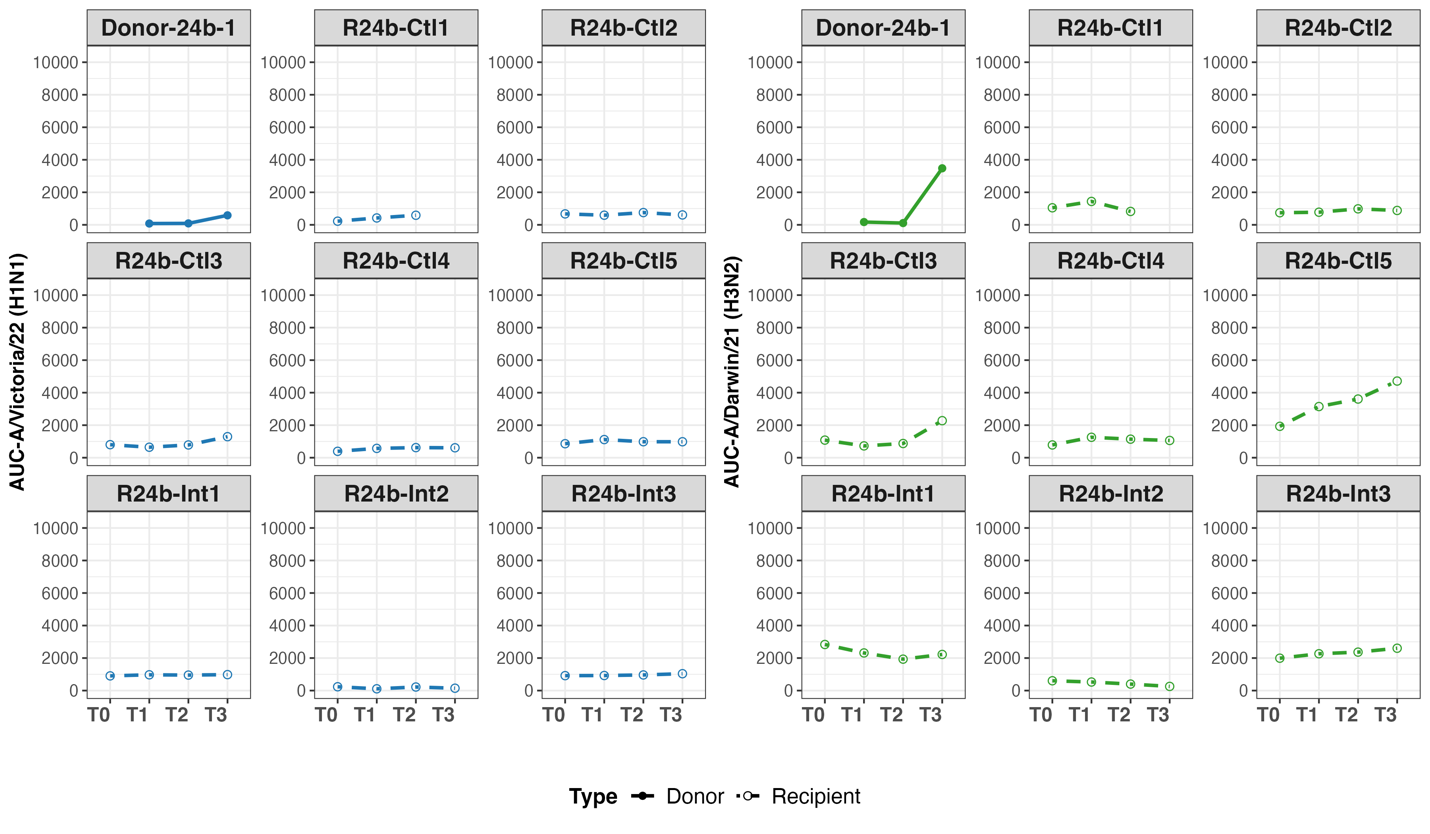

### S5 Fig. ELISA Area Under the Curve (AUC) over time for Donors and Recipients in Cohort 24b

ELISA AUC values are shown for Donors (solid lines, filled circles) and Recipients (dot-dashed lines, open circles) at four time points: T0 (Recipient screening), T1 (Admission), T2 (Discharge), and T3 (Follow-up). Virus targets are color-coded: A/Victoria/4897/22 (H1N1, blue) and A/Darwin/6/21 (H3N2, green). Each facet represents an individual study participant.
